# The contribution of animal antibiotic use to antibiotic resistance in humans: Panel evidence from Denmark

**DOI:** 10.1101/2024.01.10.24301091

**Authors:** Eve Emes, Dagim Belay, Gwenan M Knight

**Affiliations:** Centre for the Mathematical Modelling of Infectious Diseases (CMMID), London School of Hygiene and Tropical Medicine; Department of Food and Resource Economics, University of Copenhagen; Antimicrobial Resistance Centre, London School of Hygiene and Tropical Medicine

**Keywords:** AMR, One Health, epidemiology, regression, Denmark

## Abstract

Antibiotic use (ABU) in animals is postulated to be a major contributor to selection of antibiotic resistance (ABR) which subsequently causes infections in human populations. However, there are few quantifications of the size of this association. Denmark, as a country with high levels of pig production and good surveillance data, is an ideal case study for exploring this association.

In this paper, we compile a dataset on ABU across several animal species and antibiotic classes, and data on the rate of antibiotic resistance (ABR) in humans across key pathogens, in Denmark over time (2010 - 2020). We run panel data regressions (fixed effects, random effects, first difference and pooled ordinary least squares) to test the association between the level of ABR in human infections and the level of ABU in animals.

Between ABR in humans and ABU in animal species, we find a positive relationship for cattle, some evidence of a positive relationship for poultry and companion animals, and a negative relationship for fish, although the latter is likely driven by confounding factors. When lagging ABU by one year, the effect of ABU in cattle and companion animals remained similar, the effect of ABU in poultry fell in size, and ABU in fish was no longer significant, perhaps due to differences in life cycle length among animal species. Additional covariates were explored, including pet populations, agricultural production and GDP per capita (at purchasing power parity), but these results were limited by the statistical power of the dataset. Under all models, animal ABU determined only a minority of the change in human ABR levels in this context with adjusted R2 ranging from 0.19 to 0.44.

This paper supports the role of animal ABU in determining human ABR levels but suggests that, despite comprising a large portion of systemwide ABU, it only explains a minority of the variation. This is likely driven in part by data limitations, and could also be due to a persistence of ABR once resistance has emerged, suggesting a significant role for socioeconomic and transmission factors in bringing ABR down to desirable levels.

**Highlights:**

- We use panel regression to explore the link between animal ABU and human ABR in Denmark
- ABU was linked to ABR in cattle, and potentially in poultry and companion animals
- However, animal ABU did not appear to be the main determinant of human ABR
- This could be due to poor data, or persistence of ABR after reaching a certain level
- Animal ABU reductions alone may be insufficient to curb ABR

## Introduction

Antibiotic resistance (ABR), the capacity of bacterial pathogens to survive in the presence of antibiotics, is considered a major and growing threat to human health worldwide(2,3). Antibiotic use (ABU) in animals is the largest form of AMU globally(4), and as such there has been international policy focus on reducing and modulating this ABU in order to lower the rate of ABR in human infections and safeguard human and animal health.

Numerous microbiological and genomic studies(5–7) support the existence of a link between animal ABU and human ABR, and there is a very strong theoretical basis for expecting ABU in animals to generate ABR in humans(8). Despite this, our knowledge of the shape and size of this relationship remains limited(8,9), and some microbiological and genomic studies fail to find consistent evidence of it(8,10–13). This has complicated implications for AMR policy decision-making in the One Health space, where policymakers need to know the likely effect of AMS interventions on the number of resistant infections in humans and animals in order to estimate the intervention benefit. Panel regression can give specific quantitative insight into this outcome, and can feed more directly into intervention design and prioritisation at the population level.

While food animals are the largest destination of global ABU(4), genomic studies have revealed significant transmission of resistomes between humans and companion animals, and have identified companion animal ABU as an important target of interventions(11). Data on food animal ABU is collected in Denmark by VetStat and is included in this dataset: few panel regressions on the determinants of human ABR have focused on companion animal ABU.

In this study we use ecological panel data regression as our methodology as we identify it as a powerful and under-explored tool for investigating the relationship between ABU and ABR(9). Use of a given antibiotic can select for resistance to that antibiotic in pathogens which colonise and infect humans, and regression models allow us to see the extent to which use of an antibiotic is statistically related to resistance to that antibiotic in pathogens of interest to us. Use of panel regression models such as fixed effects and random effects allow for differences in the way that ABU relates to ABR among different pathogens and different drugs, given that different pathogens and different antibiotics behave differently from each other.

Ecological panel regression has been used in a number of contexts to investigate the relationship between animal ABU and human ABR, although to our knowledge this is the first study to apply it to detailed data from Denmark specifically. Rahman and Hollis(14) found that, across a panel of European countries, ABU in food animals and in humans were independently and causally related to the rate of ABR in both humans and animals. Adda(15) found that, in the United States, ABU in humans and animals both contributed to the rate of ABR in human infections, with human ABU being a greater contributor and with more recently-introduced antibiotics having a greater effect. More recently, Allel *et al*.(16) found that, across a range of countries, ABU in animals and humans contributed to the rate of ABR in infections by critical priority pathogens in humans. Zhang *et al*.(17) found a positive relationship between human ABU and the rate of fluoroquinolone resistance in *E. coli* and *P. aeruginosa* in Europe, and a negative relationship between animal ABU and fluoroquinolone resistance in *P. aeruginosa*. These studies illustrate the potential use of ecological panel regression in quantifying the relationship between animal ABU and human ABR.

Studies have also used panel regression methods to investigate the role of non-ABU factors, including socioeconomic variables and medical staffing, in determining ABR rates in humans. Collignon *et al*.(18) found that, across a range of countries and for a set of key drug-pathogen combinations, indices of infrastructure and governance were inversely related to the rate of ABR in human infections, even when human ABU was not. Zhang *et al*.(17) found that medical and veterinary staffing numbers were negatively related to the rate of fluoroquinolone resistance in *E. coli* and *P. aeruginosa* across European countries. Allel *et al*.(16) also found links between socioeconomic, demographic, political and environmental factors and human ABR across a range of countries. ABR can therefore be seen not as a purely biological problem but as a public health phenomenon which is jointly determined by biological and socioeconomic factors.

Denmark is a strong case study to investigate the relationship between animal ABU and human ABR due to the comprehensiveness of its ABR surveillance infrastructure across the One Health space, with the Danish Integrated Antimicrobial Resistance Monitoring and Research Programme (DANMAP)(19) and VetStat(20,21) tracking ABU and ABR in humans and animals. The human ABR data available through DanMap also focuses on *Campylobacter* and *Salmonella* species, which are key foodborne pathogens of relevance to human health(22). Because these pathogens are often transferred from food animals(19,23,24), they are also more likely candidates to give insight into the relationship between animal ABU and human ABR.

Denmark is considered a world leader in preventing and managing ABR from a One Health perspective: use of antibiotics in animal health has been low and consistent since 2000, and agricultural growth promoters have been phased out since then(25,26).

Denmark is also considered a world leader in agricultural AMS(27): since 1995, a series of policies has been implemented aiming to regulate and limit the use of antibiotics in animals, including bans on agricultural growth promoters from 1998(28). Animal antibiotics are sold on a prescription basis and veterinarians may not profit from their sale(27). The 2010 Yellow Card Initiative(29) places quantitative restrictions on use of antibiotics in food animal production, and has been adjusted since then to place different weights on various antibiotics depending on AMS priorities. Finally, as a country with a large amount of food animal production, particularly of pork(30), Denmark represents a strong case study for investigating the relationship between ABU in animals and the rate of ABR in human infections.

ABU may also have a delayed effect on the rate of ABR(14), especially in food animal production, where antibiotics used at the beginning of production cycles may take time to pass into the human population. Understanding the role of lagged ABU can help us to understand these transmission mechanisms.

Based on these considerations, this paper aims to investigate if ABR in human infections in Denmark is linked to the quantity of antibiotics used in animals, and to quantify that link. And, if a relationship is observed, to determine whether or not it varies among animal species. After addressing these questions, we will explore the shape and nonlinearity of that relationship. Finally, we will investigate whether antibiotic use in previous periods is linked to the rate of ABR, and how strong this link is compared with that of same-period ABR, as well as exploring the role of other covariates including GDP per capita and animal populations. These covariates will help to account for changing socioeconomic conditions which could influence the relationship between ABU and ABR, as well as potential relationships between populations of, and therefore use of antibiotics in, different animal types.

## Materials and Methods

### Data

Data on the rate of ABR in humans was sourced from DanMap(19), the Danish Integrated Antimicrobial Resistance Monitoring and Research Programme. DanMap makes publicly available a repository of data on ABR indicators and zoonotic bacteria in humans, livestock and companion animals in Denmark, drawing on routine surveillance across primary and secondary healthcare, veterinary surveillance and prevalence surveys from livestock animals. In humans, data coverage is high - representing a near complete proportion of all microbiological analyses. The source of the bacterial sample depends on the species, ranging from bloodstream infections to colonisation samples. In the following we term “human ABR” to be the proportion of isolates for a certain bacterial species collected by DANMAP in routine surveillance (often only the first isolate from a patient per year) that were tested and found to be resistant to the antibiotic being considered(19).

Data on the use of antibiotics in food and companion animals was sourced from VETSTAT(20), a database which records all prescription drugs sold for animal use in Denmark. In this dataset, we refer to the total amount of each antibiotic prescribed for use in each animal type, by kg of active compound, each year.

### Variables

We cleaned and compiled the data into a panel at the {*year*, *drug-pathogen*} level. Drug-pathogen refers to the observed rate of resistance of isolates of a particular bacteria species (pathogen) to a specific class of antibiotic (drug). For example, the rate of resistance of Salmonella typhimurium to tetracyclines represents one drug-pathogen pair.

For each year, and each drug-pathogen pair, we used data on:

- The portion of human bacterial isolates which were resistant to various antibiotics, from routine healthcare surveillance, from 2010 to 2021.
- The total use of antibiotics in kg in several livestock animal types, and for companion animals, from 2010 to 2020

Antibiotics here were sorted at the class level. While the use of antibiotics was recorded by antibiotic class, the resistance dataset recorded resistance against several individual drugs. For this reason, we grouped drugs into classes(31), using the average rate of resistance against all drugs from each antibiotic class. For more detail on the classification of antibiotics in this study, see Appendix 1. The pathogens covered by the dataset include *Campylobacter coli*, *Campylobacter jejuni*, *Escherichia coli*, *Salmonella derby*, *Salmonella enteritidis*, *Salmonella infantis* and *Salmonella typhimurium*.

The animal types included in our study were: cattle, sheep and goats, pigs, poultry, fish, and companion animals.

### Statistical methods

We first cleaned the raw datasets by extracting relevant data, standardising the classification of antibiotics across the two datasets, aggregating data into a {*year*, *drug-pathogen*} panel, and merging the two datasets. We explored the data coverage and completeness across humans and animals and across the different years and drug-pathogen pairs covered.

We generated summary statistics on the use of antibiotics by animal species and class over time, as well as on the rate of resistance in human isolates over time (by drug-pathogen combination).

For our regression analysis, we used fixed effects, random effects, first difference, and pooled ordinary least squares (POLS) regressions. A Durbin-Wu-Hausman test(32) was used to determine whether or not random effects models should be included.

First, we performed multivariate regression analysis, regressing human ABR against ABU in each animal species together. This gives us our main regression models (below)

Fixed effects

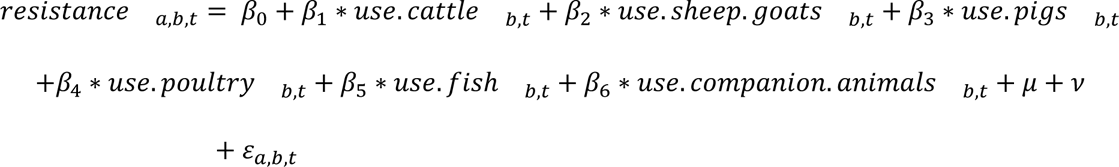

Random effects and POLS

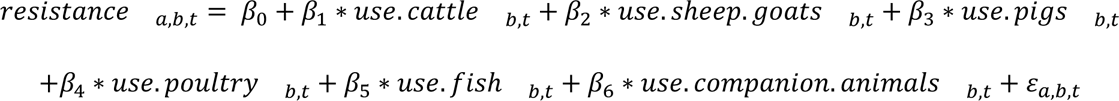

First difference

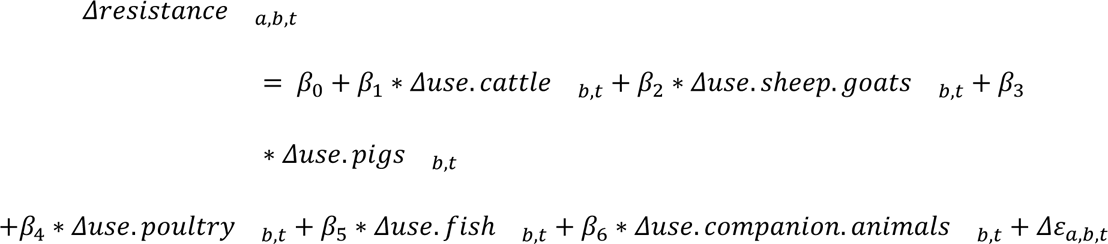

Where:

- β _0_ is the intercept and β _1−6_ are the regression coefficients,
- Δ refers to the change in a variable between year *t* − 1 and year *t*,
- *resistance _a,b,t_* is the portion of tested human isolates from pathogen *a* which were resistant to antibiotic *b* in year *t*,
- *use*. *animal* _*b,t*_ is the quantity of antibiotic *b* used in each given animal type in year *t*
- μ and *v* are the year and drug-pathogen fixed effects (fixed effects model only), and
- ε_*a,b,t*_ is the error term

That is, use of antibiotic *b* in each animal in year *t* may affect the rate of resistance of tested human isolates of pathogen *a* to antibiotic *b* in year *t*. Random effects, fixed effects and first difference models allow this relationship to vary among drug-pathogen pairs. A β coefficient of 1 means that an increase in ABU in a given animal type of 1kg per year is associated with a one percent point increase in the portion of tested human isolates which were resistant to that antibiotic class.

After this, we performed univariate analyses, regressing human ABR against ABU in each livestock species individually.

Following this, we reran the multivariate specifications against ABU lagged by one year. Finally, we reran the univariate specifications while including a quadratic term, to explore nonlinearities.

Finally, we reran the main univariate and multivariate specifications with the addition of key covariates. Namely: GDP per capita (at purchasing power parity), population of each livestock species, and pet ownership, over time. GDP per capita was included due to the potential role of socioeconomic covariates discussed earlier(16–18). Animal populations were included because, while the population of each animal type is likely positively related to total ABU in that animal type, populations of each animal may also be negatively related to each other, due to substitutability between different meat types. For example, if cow and sheep meat have a negative cross-elasticity of demand, then an increase in cow production (and therefore an increase in ABU in cows) may engender a fall in the population of (and therefore ABU in) sheep, while simultaneously resulting in an increase in human ABR. This could create the erroneous impression that the fall in ABU in sheep caused a rise in human ABR, creating the appearance of a negative relationship between sheep ABU and human ABR.

Data on GDP per capita (PPP) was sourced from World Bank Open Data(33), and data on animal populations came from Statistics Denmark(34).

## Results

### Summary statistics

Our (combined DanMap - VetStat) dataset had 62 different drug-pathogen combinations across 7 bacterial species and 11 antibiotic classes. Data on ABR covered 2010 - 2021 and data on ABU covered 2010 - 2020 (11 years). We used 7 ABU and ABR variables in this investigation (ABR in humans, and ABU in 6 different animal types). Across 7 variables, 7 pathogen types, 11 antibiotic classes, and 11 years, a complete dataset would have 5929 observations across 847 year-drug-pathogen combinations.

In our dataset, we had:

- 893 non-NA observations (15.1% completeness)
- 149 year-drug-pathogen combinations with data on human ABR (17.6% completeness)
- 124 year-drug-pathogen with data on animal ABU (14.6% completeness)
- 48 year-drug-pathogens with data on both human ABR and animal ABU (5.7% completeness)

We can thus see that, while a complete dataset would have had a very large number of datapoints, missingness greatly reduced our statistical power. Not only that, but the very low overlap between year-drug-pathogen combinations with data on human ABR and animal ABU meant that we were effectively left with only 48 observations, creating statistical power issues especially when (year and drug-pathogen) fixed effects or covariates are introduced. We were nevertheless able to get significant results in certain specifications, and the inclusion of different models (fixed effects, random effects, first difference, and POLS) helped to tease out relationships.

As we can see from the summary statistics (Fig. 1), total use of sulfonamides in animals has fallen slowly and consistently over the study period, and use of tetracyclines has fallen considerably. The latter is largely driven by use in pigs (which comprises the bulk of tetracycline use), in which there was a sharp decline from 2015-2018, although declines also occurred in poultry and sheep and goats during that time (Fig. 2). There have also been noticeable falls in the use of sulfonamides in fish from 2013 to 2017 (though at < 5% of total use), and in the use of tetracyclines in sheep and goats from 2010 to 2012 (though always at < 1% of total use) (Fig. 2). By contrast, use of tetracyclines in poultry rose from 2012 to 2015, and use of sulfonamides in poultry spiked in 2015 (Fig. 2). Note that the total quantity of antibiotics used varied considerably by animal type. Pigs accounted for the most by far (78% of all use recorded in the dataset), followed by cattle (7.3%), then poultry (1.4%), then companion animals (0.60%) and fish (0.49%), with sheep and goats (0.022%) accounting for the least total ABU.

**Fig. 1.**
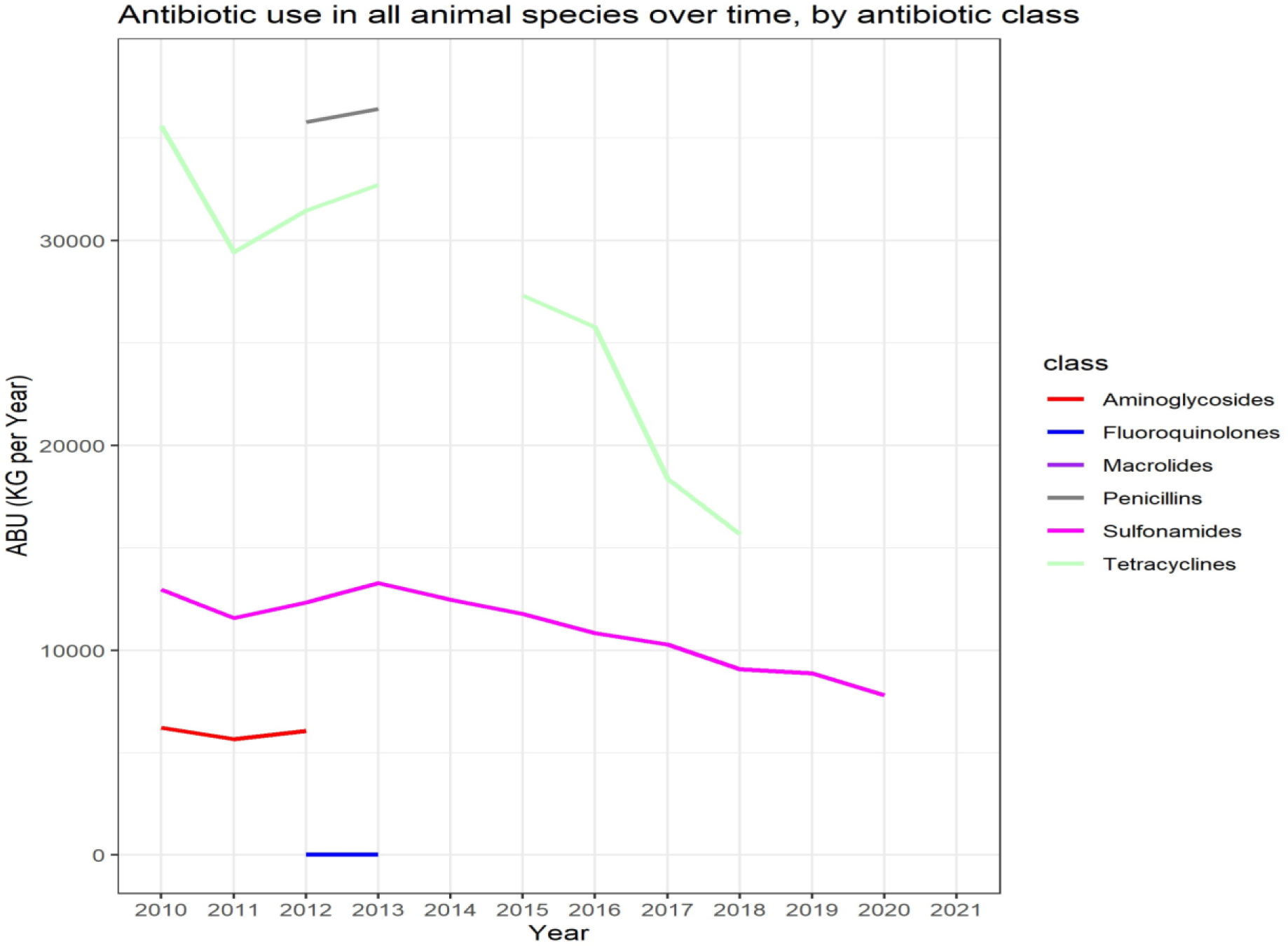
Antibiotic use (kg per year) in all animal types over time, by antibiotic class.

**Fig. 2.**
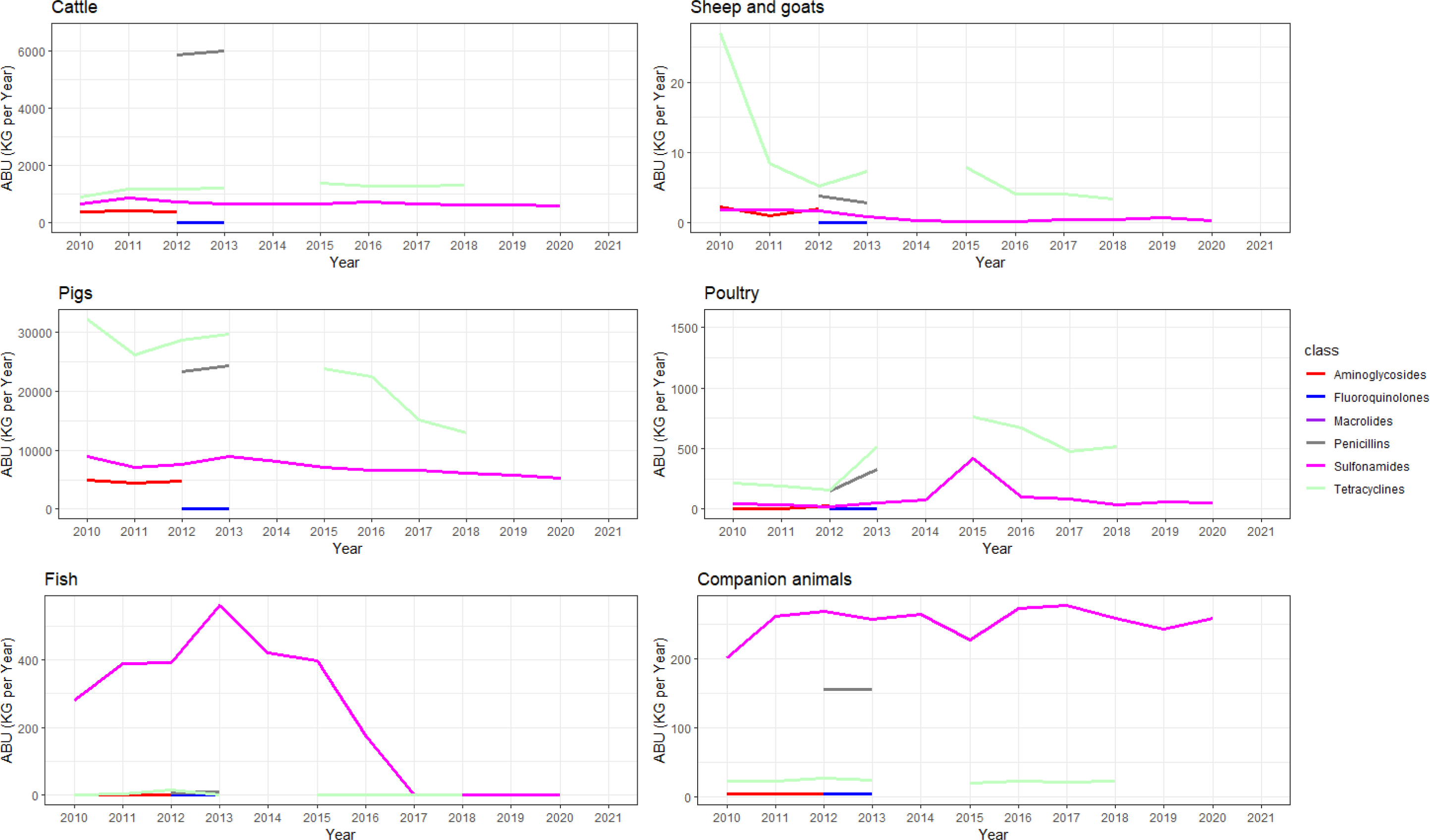
Antibiotic use (kg per year) over time in each livestock species, by antibiotic class.

The rate of ABR in humans has remained relatively consistent during the study period (Fig. 3), or risen for some of the drug-pathogen pairs with the highest observed rate of resistance, with resistance of *C. jejuni* and *S. typhimurium* to certain key antibiotics being considerably higher than resistance in other drug-pathogen combinations. In particular, resistance to tetracyclines nearly doubled in these pathogens from 2010 to 2018.

**Fig. 3.**
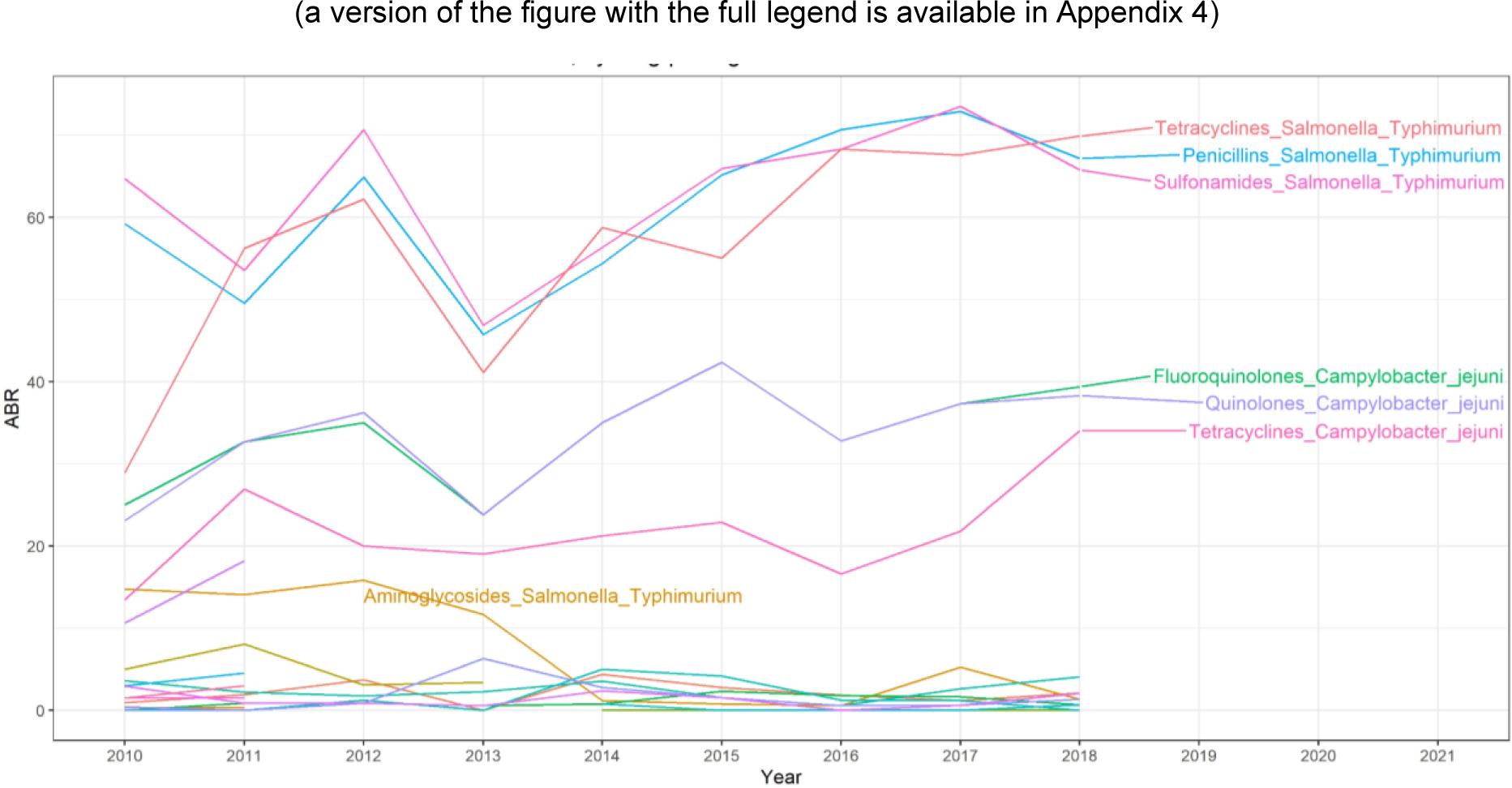
Rate of ABR in humans over time in Denmark, by drug-pathogen combination.

### Multivariate specifications

We used a Durbin-Wu-Hausman test(32) to determine whether random effects should be used. We failed to reject the null hypothesis, indicating that the random effects model was more efficient and no less consistent than fixed effects, and so both fixed and random effects models were included.

After running the multivariate specifications (Table 1), ABU in cattle was positively associated with ABR in humans in the random effects and first difference specifications. ABU in poultry was positively associated with human ABR in the POLS regression. ABU in fish was negatively associated with human ABR in the random effects and first difference specifications, and ABU in companion animals was strongly positively associated with human ABR in the POLS specification only. All of the specifications were jointly significant, except for the fixed effects regression (as measured by the F statistic). Of the three significant specifications, the adjusted R^2^ ranged between 0.188 and 0.443. ABU in pigs was not associated with ABR in humans in any model.

**Table 1.**
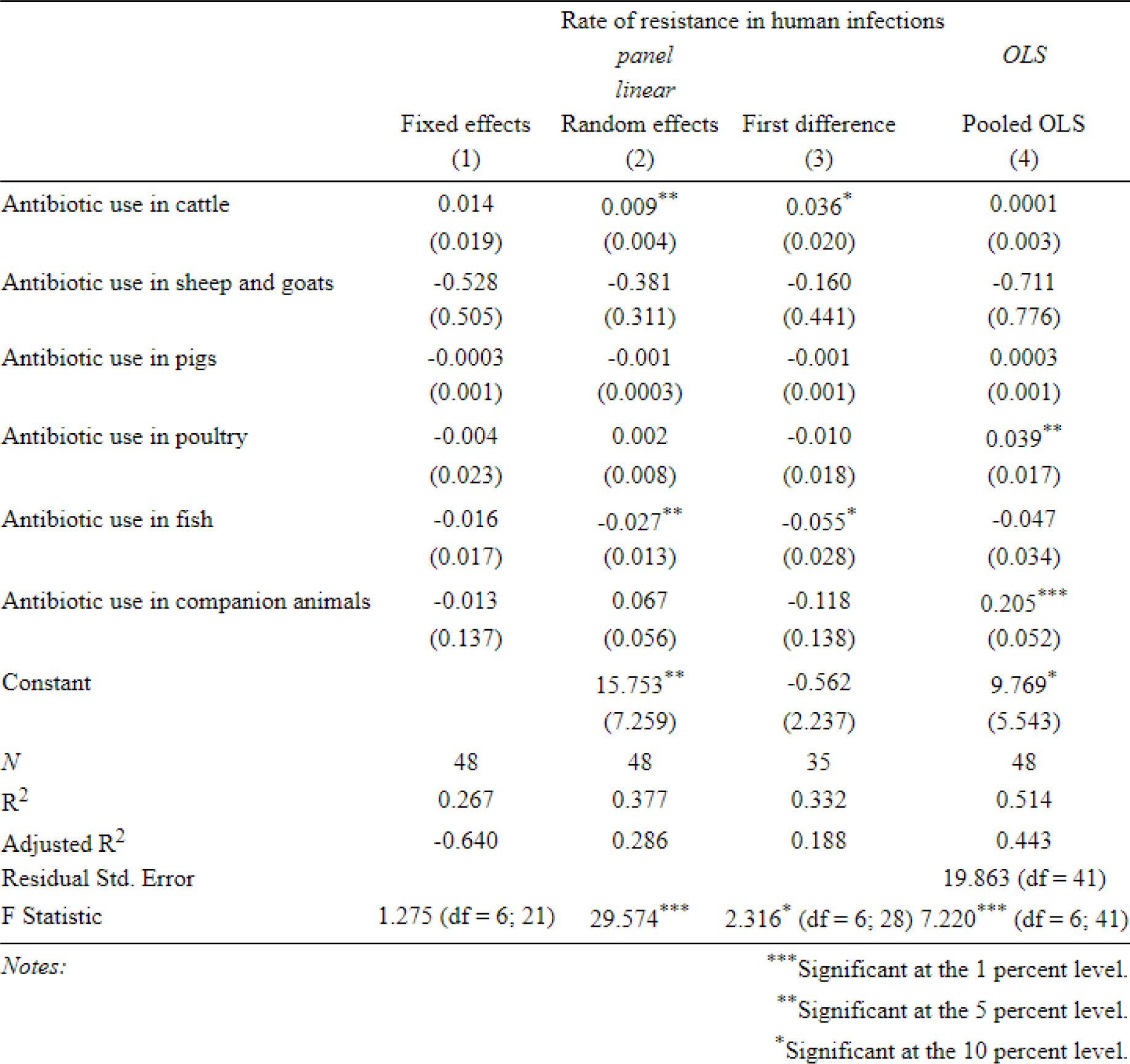
multivariate specifications.

|  | Rate of resistance in human infections |  |  | *OLS* |
| --- | --- | --- | --- | --- |
|  | Fixed effects | *panel*<br>*linear*<br>Random effects | First difference | Pooled OLS |
|  | (1) | (2) | (3) | (4) |
| Antibiotic use in cattle | 0.014<br>(0.019) | 0.009**<br>(0.004) | 0.036*<br>(0.020) | 0.0001<br>(0.003) |
| Antibiotic use in sheep and goats | -0.528<br>(0.505) | -0.381<br>(0.311) | -0.160<br>(0.441) | -0.711<br>(0.776) |
| Antibiotic use in pigs | -0.0003<br>(0.001) | -0.001<br>(0.0003) | -0.001<br>(0.001) | 0.0003<br>(0.001) |
| Antibiotic use in poultry | -0.004<br>(0.023) | 0.002<br>(0.008) | -0.010<br>(0.018) | 0.039**<br>(0.017) |
| Antibiotic use in fish | -0.016<br>(0.017) | -0.027**<br>(0.013) | -0.055*<br>(0.028) | -0.047<br>(0.034) |
| Antibiotic use in companion animals | -0.013<br>(0.137) | 0.067<br>(0.056) | -0.118<br>(0.138) | 0.205***<br>(0.052) |
| Constant |  | 15.753**<br>(7.259) | -0.562<br>(2.237) | 9.769*<br>(5.543) |
| *N* | 48 | 48 | 35 | 48 |
| *R*<sup>2</sup> | 0.267 | 0.377 | 0.332 | 0.514 |
| Adjusted *R*<sup>2</sup> | -0.640 | 0.286 | 0.188 | 0.443 |
| Residual Std. Error |  |  |  | 19.863 (df = 41) |
| F Statistic | 1.275 (df = 6; 21) | 29.574*** | 2.316* (df = 6; 28) | 7.220*** (df = 6; 41) |

Notes:
\*\*\*Significant at the 1 percent level.
\*\*Significant at the 5 percent level.
\*Significant at the 10 percent level.

### Univariate specifications

After running the univariate specifications (Table 2), ABU in cattle was positively associated with human ABR in the random effects, first difference, and POLS regressions (Table 2.1). ABU in sheep and goats was negatively associated with human ABR in the fixed effects, random effects and first difference specifications (Table 2.2). ABU in pigs was negatively associated with human ABR in the random effects and first difference specifications (Table 2.3). ABU in poultry was positively associated with human ABR in the random effects and POLS specifications (Table 2.4). ABU in fish was negatively associated with human ABR in the random effects specification, but positively associated with human ABR in the POLS specification (Table 2.5). Finally, ABU in companion animals was positively associated with human ABR in the random effects and POLS specifications.

**Table 2.** Univariate specifications for each animal type.

**Table 2.1.** univariate regressions (cattle)

|  | Rate of resistance in human infections |  |  | OLS |
| --- | --- | --- | --- | --- |
|  | Fixed effects | Random effects | First difference | Pooled OLS |
|  | (1) | (2) | (3) | (4) |
| Antibiotic use in cattle | 0.017<br>(0.011) | 0.012***<br>(0.004) | 0.029**<br>(0.013) | 0.007***<br>(0.003) |
| Constant |  | 8.105<br>(8.150) | 0.505<br>(2.005) | 24.022***<br>(4.520) |
| N | 48 | 48 | 35 | 48 |
| R<sup>2</sup> | 0.085 | 0.114 | 0.134 | 0.153 |
| Adjusted R<sup>2</sup> | -0.655 | 0.095 | 0.108 | 0.135 |
| Residual Std. Error |  |  |  | 24.747 (df = 46) |
| F Statistic | 2.407 (df = 1; 26) | 8.973*** | 5.120** (df = 1; 33) | 8.323*** (df = 1; 46) |
| Notes: |  |  |  | *** Significant at the 1 percent level.<br>** Significant at the 5 percent level.<br>* Significant at the 10 percent level. |

**Table 2.2.**
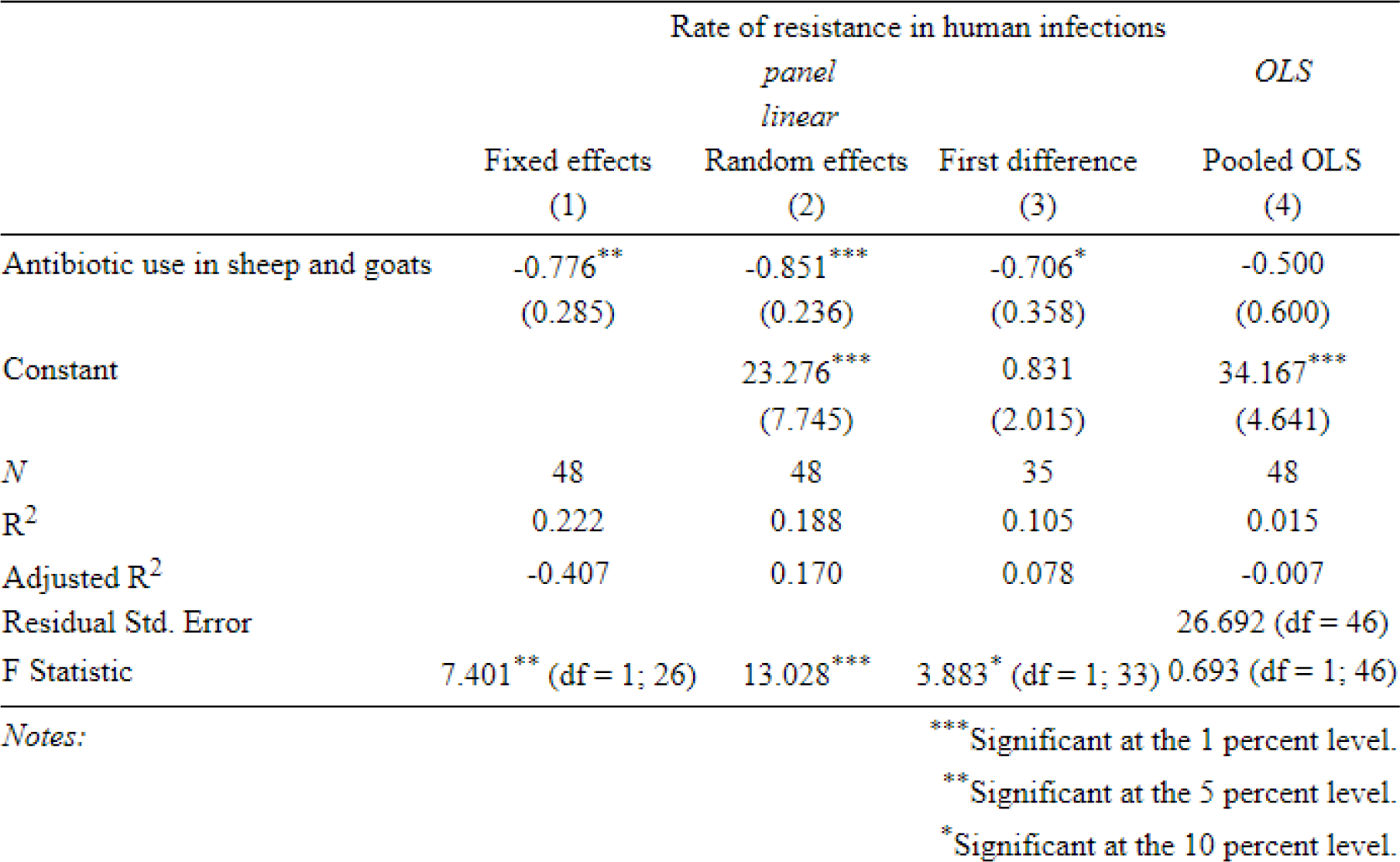
univariate regressions (sheep and goats)

|  | Rate of resistance in human infections |  |  | OLS |
| --- | --- | --- | --- | --- |
|  | Fixed effects | Random effects | First difference | Pooled OLS |
|  | (1) | (2) | (3) | (4) |
| Antibiotic use in sheep and goats | -0.776**<br>(0.285) | -0.851***<br>(0.236) | -0.706*<br>(0.358) | -0.500<br>(0.600) |
| Constant |  | 23.276***<br>(7.745) | 0.831<br>(2.015) | 34.167***<br>(4.641) |
| N | 48 | 48 | 35 | 48 |
| R<sup>2</sup> | 0.222 | 0.188 | 0.105 | 0.015 |
| Adjusted R<sup>2</sup> | -0.407 | 0.170 | 0.078 | -0.007 |
| Residual Std. Error |  |  |  | 26.692 (df = 46) |
| F Statistic | 7.401** (df = 1; 26) | 13.028*** | 3.883* (df = 1; 33) | 0.693 (df = 1; 46) |
| Notes: |  |  |  | *** Significant at the 1 percent level.<br>** Significant at the 5 percent level.<br>* Significant at the 10 percent level. |

**Table 2.3.** univariate regressions (pigs)

|  | Rate of resistance in human infections |  |  |  |
| --- | --- | --- | --- | --- |
|  | *panel*<br>*linear* |  |  | *OLS* |
|  | Fixed effects<br>(1) | Random effects<br>(2) | First difference<br>(3) | Pooled OLS<br>(4) |
| Antibiotic use in pigs | -0.001<br>(0.0004) | -0.001***<br>(0.0003) | -0.001**<br>(0.001) | 0.0004<br>(0.0004) |
| Constant |  | 31.196***<br>(8.501) | 0.185<br>(2.089) | 26.382***<br>(6.240) |
| *N* | 48 | 48 | 35 | 48 |
| *R*<sup>2</sup> | 0.075 | 0.174 | 0.125 | 0.028 |
| Adjusted *R*<sup>2</sup> | -0.673 | 0.156 | 0.099 | 0.006 |
| Residual Std. Error |  |  |  | 26.519 (df = 46) |
| F Statistic | 2.095 (df = 1; 26) | 11.793*** | 4.730** (df = 1; 33) | 1.306 (df = 1; 46) |
| *Notes:* |  |  | ***Significant at the 1 percent level. |  |
|  |  |  | **Significant at the 5 percent level. |  |
|  |  |  | *Significant at the 10 percent level. |  |

**Table 2.4.** univariate regressions (poultry)

|  | Rate of resistance in human infections |  |  |  |
| --- | --- | --- | --- | --- |
|  | *panel*<br>*linear* |  |  | *OLS* |
|  | Fixed effects<br>(1) | Random effects<br>(2) | First difference<br>(3) | Pooled OLS<br>(4) |
| Antibiotic use in poultry | 0.012<br>(0.015) | 0.015*<br>(0.009) | -0.003<br>(0.013) | 0.036**<br>(0.016) |
| Constant |  | 18.133**<br>(7.405) | 2.342<br>(2.027) | 24.714***<br>(4.896) |
| *N* | 48 | 48 | 35 | 48 |
| *R*<sup>2</sup> | 0.025 | 0.015 | 0.001 | 0.100 |
| Adjusted *R*<sup>2</sup> | -0.762 | -0.006 | -0.029 | 0.081 |
| Residual Std. Error |  |  |  | 25.511 (df = 46) |
| F Statistic | 0.677 (df = 1; 26) | 2.917* | 0.042 (df = 1; 33) | 5.118** (df = 1; 46) |
| *Notes:* |  |  | ***Significant at the 1 percent level. |  |
|  |  |  | **Significant at the 5 percent level. |  |
|  |  |  | *Significant at the 10 percent level. |  |

**Table 2.5.**
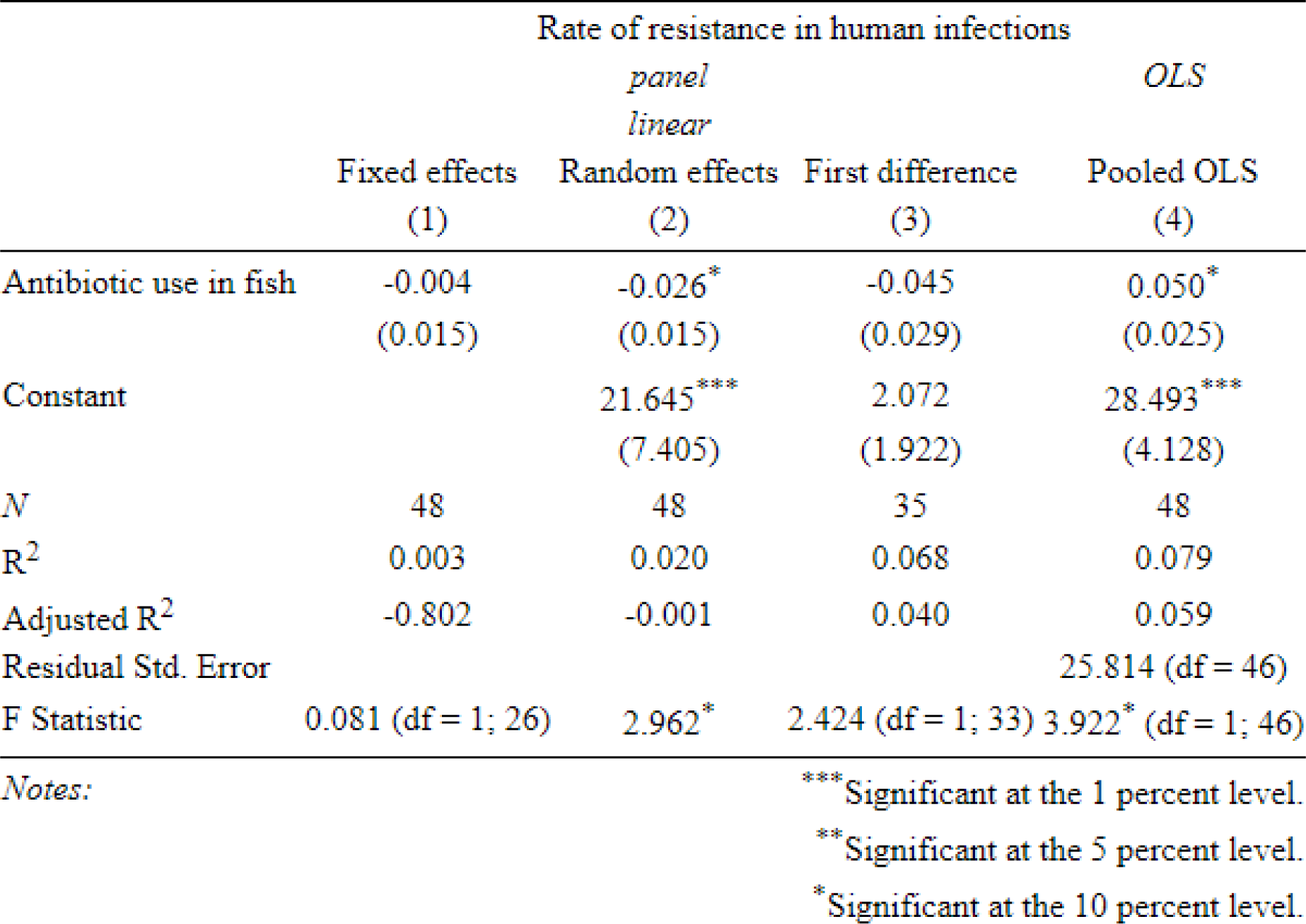
univariate regressions (fish)

**Table 2.6.**
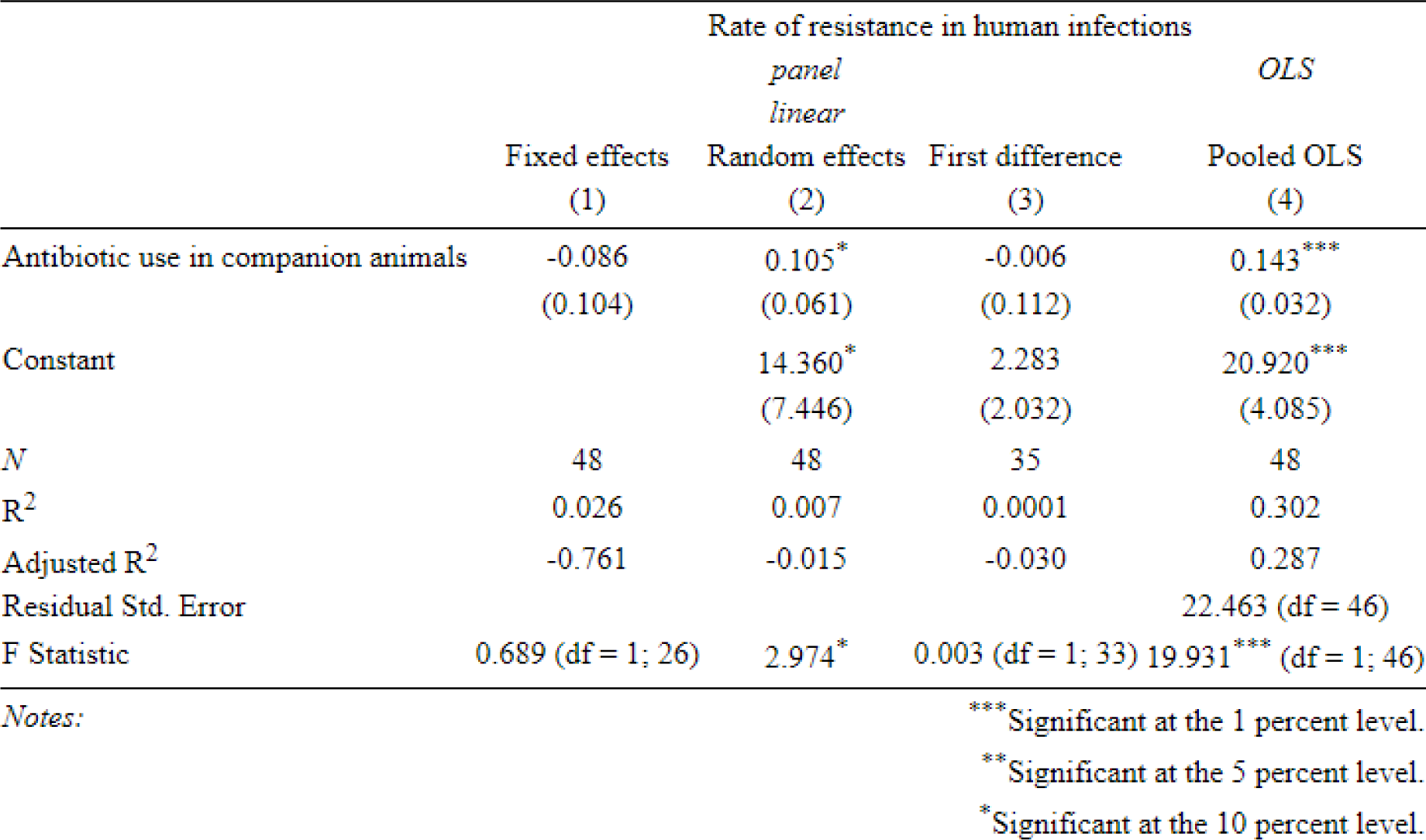
univariate regressions (companion animals)

### Lagged independent variable

When lagging animal ABU by one year (Table 3), ABU in cattle was still positively associated with ABR in humans in the random effects and first difference specifications, with the effect size remaining similar to the same-period model. ABU in poultry remained positively associated with human ABR in the POLS regression, with the effect size falling. ABU in fish was no longer associated with human ABR; and ABU in companion animals remained positively associated with human ABR in the POLS specification, with the effect size remaining similar. ABU in pigs remained without an association.

**Table 3.** multivariate specifications (independent variables lagged by one year)

|  | Rate of resistance in human infections |  |  | OLS |
| --- | --- | --- | --- | --- |
|  | Fixed effects | Random effects | First difference | Pooled OLS |
|  | (1) | (2) | (3) | (4) |
| Lagged antibiotic use in cattle | 0.008<br>(0.013) | 0.009**<br>(0.004) | 0.030**<br>(0.013) | 0.0001<br>(0.003) |
| Lagged antibiotic use in sheep and goats | 0.635<br>(0.451) | 0.382<br>(0.367) | 0.687<br>(0.435) | -0.278<br>(0.737) |
| Lagged antibiotic use in pigs | -0.001<br>(0.001) | -0.0004<br>(0.0004) | -0.0004<br>(0.001) | 0.001<br>(0.001) |
| Lagged antibiotic use in poultry | 0.009<br>(0.008) | 0.003<br>(0.006) | -0.006<br>(0.006) | 0.019**<br>(0.009) |
| Lagged antibiotic use in fish | -0.006<br>(0.017) | 0.007<br>(0.014) | 0.018<br>(0.028) | -0.002<br>(0.033) |
| Lagged antibiotic use in companion animals | 0.017<br>(0.126) | 0.081<br>(0.063) | 0.076<br>(0.125) | 0.163***<br>(0.051) |
| Constant |  | 10.012<br>(7.944) | 0.684<br>(2.144) | 8.974<br>(5.573) |
| N | 45 | 45 | 32 | 45 |
| R<sup>2</sup> | 0.253 | 0.148 | 0.291 | 0.508 |
| Adjusted R<sup>2</sup> | -0.826 | 0.014 | 0.121 | 0.431 |
| Residual Std. Error |  |  |  | 19.217 (df = 38) |
| F Statistic | 1.017 (df = 6; 18) | 11.799* | 1.714 (df = 6; 25) | 6.550*** (df = 6; 38) |

Notes:
\*\*\*Significant at the 1 percent level.
\*\*Significant at the 5 percent level.
\*Significant at the 10 percent level.

### Additional specifications

After this, we reran the univariate specifications with the addition of a quadratic term. However, we were not able to identify any consistent trends (Appendix 2).

Finally, we reran the main univariate and multivariate specifications with the addition of key covariates (GDP per capita at purchasing power parity and animal populations). For the multivariate specification, populations of all animal types were included, while for the univariate specifications only the population of only one animal type at a time was included. With the addition of these covariates, the multivariate models could not be estimated due to a lack of data.

For the univariate models, we had to drop covariates in some cases due to multicollinearity or a lack of data (especially for fish, where we only had data on fisheries production since 2017) (Appendix 3). Animal populations were never significantly related to human ABR. GDP per capita (PPP) was positively related to human ABR in some specifications, although this may simply be due to the fact that Denmark’s per-person income has consistently increased during the study period, with human ABR rising somewhat as well.

Controlling for animal population and GDP per capita (PPP), ABU in companion animals remained positively related to human ABR in the random effects and POLS models and ABU in cattle was positively related to human ABR in the POLS model (Appendix 3).

## Discussion

### Findings and interpretation

Across our univariate and multivariate specifications, we found evidence that ABU in cattle, poultry and companion animals was positively associated with human ABR. The evidence for cattle was the most consistent, and the effect size was greatest for companion animals. The effect size varied greatly between animal types, although this may be simply due to great differences in the volume of antibiotics used in each animal type.

ABU in sheep and goats, as well as in pigs, was negatively associated with human ABR in some univariate specifications but not in the multivariate specifications. ABU in fish was negatively associated with human ABR in some multivariate specifications, and had an indeterminate relationship to human ABR in the univariate specifications. However, ABU in fish comprised such a small component of total ABU that we cannot infer causality from that result. This may instead be due to a fall in the use of sulfonamides in fish during the study period concurrent with stable or increasing overall levels of ABR in humans driven by other factors.

When lagging antibiotic use by one year, the effects identified in the same-period models remained similar for animals with longer life-cycles (companion animals and cattle). For animals with shorter life cycles the effect either fell in size (poultry) or was no longer significant (fish). We did not identify any consistent trends when rerunning the univariate specifications with the addition of a quadratic term.

While the multivariate models could not be run with the inclusion of additional covariates, running the univariate models while controlling for animal populations and GDP per capita (PPP) revealed a positive relationship between human ABR and ABU in companion animals and, to a lesser extent, in cattle.

In the multivariate specifications which were jointly significant, the adjusted R^2^ ranged between 0.188 and 0.443. This suggests that ABU in animal health does explain a significant portion of variation in human ABR but, despite accounting for the majority of systemwide ABU (and two thirds of all ABU globally(35)), is not responsible for the majority of this variation. The effect size observed varied considerably between different animal species, though this may partially reflect large differences in total production and total ABU across different animal types.

It is counterintuitive that negative relationships were observed between human ABR and ABU in some animal species. In the case of pigs, sheep and goats, this may be due to a negative cross-elasticity of demand between consumption of cattle and consumption of pork, lamb and mutton. That is to say, if production of (and therefore use of antibiotics in) pigs, sheep and goats is negatively related to production of (and therefore use of antibiotics in) cattle, then the positive relationship between ABU in cattle and human ABR may create the impression of a negative relationship between ABU in pigs, sheep and goats and ABR in humans in the univariate specifications. This would also explain why those negative relationships were not observed in the multivariate specifications.

While ABU in pigs accounted for the considerable majority of animal ABU during the study period, it was not associated with human ABR in any of the multivariate specifications. This runs counter to the hypothesis that total volume of animal ABU correlates to the rate of human ABR. Our results ABU in fish was negatively associated with human ABR even in the multivariate specifications. However, this may be due to the significant reduction in the use of sulfonamides in fish production (Fig. 2) concurrent with a generally stable or slightly increasing rate of human ABR (Fig. 3). ABU in fish accounted for such a small portion of total ABU that concurrent trends such as this may drive statistical associations more than an underlying causality.

### Limitations

A major limitation of our analysis was the suitability of publically open access available data. While considerable data on ABU and ABR were available, the overlap of years and antibiotic classes covered by the ABU and ABR datasets was limited, meaning that our statistical power was similarly limited. This prevented more detailed investigations of the shape of the ABU-ABR relationship, into the role of other covariates, or on what relationships could be observed for specific antibiotic classes and specific bacterial pathogens.

There was also relatively little change in the use of certain antibiotics in certain animals during the study period, and even where large relative changes were observed, the starting level of ABU is low compared with other contexts. Both animal ABU and human ABR in Denmark have been closely managed since some years before this dataset begins(25,26), meaning that we might not expect these changes to greatly influence human ABR.

An important limitation with this kind of investigation is the notion that, while use of antibiotics by humans (in both humans and animals) is generally agreed to have created the ongoing ABR pandemic(8), this does not necessarily mean that reductions in ABU will result in reductions in ABR. Allel *et al*.(16) also emphasise that ABU reduction alone is unlikely to bring down the rate of ABR in human infections significantly. This ‘stickiness’ of ABR, especially in a context like Denmark where rates of resistance are already relatively low and stable, means that associations between ABU and ABR may not be statistically significant, or may be obscured by factors such as negative cross-elasticity of demand among meat types. Similarly, in cases like the use of sulfonamides in fish, large reductions in certain types of ABU combined with stable or increasing rates of human ABR can generate negative statistical associations between ABU and ABR when a causal association may not exist.

The dataset also only covered resistance in human isolates of *Campylobacter* and *Salmonella* species, and *E. coli*. While these are important foodborne pathogens, they are not reflective of the total human ABR burden, and links between animal ABU and human ABR may have been observable for other pathogens had we had data on them.

Finally, while the Durbin-Wu-Hausman test suggested that random effects models were consistent, we may have failed to reject the null hypothesis of this test in part due to limited statistical power. If our covariates (animal ABU) were indeed determined in large part by time-invariant unobservables, then the results of our random effects models would become inconsistent.

### Implications for research, policy, and practice

In this study we found some evidence of animal ABU contributing to human ABR in Denmark, consistent with other ecological regression studies. Allel *et al*.(16) found this to be the case across a number of countries, for certain drug-pathogen combinations. Rahman and Hollis(14) found more consistent evidence of this across European countries for a range of drug-pathogen combinations.

While we did find some evidence of association, animal ABU did not explain the majority of variation in human ABR and results for some livestock species were not consistently significant. This could suggest, as Adda(15) found in the United States, that while animal ABU had some influence on human ABR, and despite animal use accounting for a large portion of total ABU, it was human ABU which was the more important determinant by far. This could also suggest that, in contexts such as Denmark where ABU in animals is limited to the minimum clinically necessary amount(25,26), the link between human ABR and animal ABU may not be pronounced. Given that resistance has plateaued or even risen for some drug-pathogen combinations in Denmark (Fig. 3), this could suggest that, once ABR reaches a certain level, ABU reductions may not be sufficient to reduce it in the short-to-medium term. This is consistent with some trends observed in our data, such as resistance in humans remaining high despite considerable reductions in use. Non-ABU factors, including transmission factors and socioeconomic factors, may be more relatively influential, especially in low-ABU contexts such as Denmark. This is consistent with the findings of Zhang *et al*.(17) and Collignon *et al*.(18), who respectively identify medical staffing and socioeconomic factors as important determinants of ABR prevalence in human infections at the population level.

Data sharing initiatives across the One Health space such as those proposed by the Quadripartite(36) will be key to future work in this area. We were able to access nationally aggregated longitudinal data from DanMap and VetStat from open access resources. However, there were limitations to this data such as differences in antibiotic class aggregation and missing timepoints that need to be addressed for optimal analysis. Moving forward, for ecological level of associations being hypothesised for ABR and to inform antibiotic stewardship across the One Health spectrum, aggregated, non-identifiable data is vital and could be shared from both human and animal sectors whilst avoiding any confidentiality issues.

Future studies should repeat these models with more comprehensive data, when available. Given the suggestion of this study, as well as of other regression studies, that ABU reductions alone may be insufficient to bring down human ABR in the short term, future studies should investigate non-ABU covariates (socioeconomic and transmission factors) which may influence human ABR and may modulate the effect of ABU on ABR, as well as looking at longer timeframes as more data become available.

## Conclusions

In this study, we used ecological regression to investigate the relationship between animal ABU and human ABR in Denmark. We found evidence of a positive relationship between ABU in cattle, poultry and companion animals and ABR in humans. A negative relationship between ABU in pigs, sheep and goats and ABR in humans was identified in the univariate specifications, but was not present in the multivariate specifications and may have been due to confounding factors. For animals with longer life cycles, lagged ABU remained related to human ABR. Our findings support the idea that animal ABU influences human ABR, but do not indicate that it is the main determinant of human ABR in Denmark. Especially in contexts such as Denmark with extensive antibiotic stewardship and antibiotic use controls, this suggests that ABU reduction alone may not be sufficient to bring down ABR rates, and that transmission-related and socioeconomic factors may play an important role in future research and policy on One Health ABR.

## Data Availability

All data produced are available online at:
DanMap (https://www.danmap.org/)
VETSTAT (https://vetstat.fvst.dk/vetstat/)

https://vetstat.fvst.dk/vetstat/

## Acknowledgements

This paper was written as part of the SEFASI consortium(1) (grant no. JPIAMR2021-182 SEFASI) under the umbrella of the JPIAMR - Joint Programming Initiative on Antimicrobial Resistance, based at the London School of Hygiene and Tropical Medicine which focuses on agriculture and antimicrobial resistance (AMR) from a One Health^1^ Perspective in England, Senegal and Denmark. The authors extend their thanks to the surveillance team at VetStat and The Danish Integrated Antimicrobial Resistance Monitoring and Research Programme (DANMAP) for curating these data and making them publicly available.

## Acronyms

ABR: Antibiotic resistance
ABU: Antibiotic use
AMR: Antimicrobial resistance
AMS: Antimicrobial stewardship
AMU: Antimicrobial use
DanMap: The Danish Integrated Antimicrobial Resistance Monitoring and Research Programme
OLS: Ordinary least squares regression
One Health: The interplay between human, animal and environmental health
POLS: Pooled OLS
SEFASI: Selecting Efficient Farm-Level Antimicrobial Stewardship Interventions from a One Health Perspective

## Author contributions

Conceptualisation: EE and GK; methodology: EE, DB and GK; software: EE; validation: EE; formal analysis: EE; investigation: EE; data curation: EE and DB; writing - original draft preparation: EE; writing - review and editing: EE, DB and GK; visualisation: EE; supervision: GK; project administration: GK; funding acquisition: EE, DB and GK. All authors have read and agreed to the published version of the manuscript.

## Institutional Review Board Statement

The study used only anonymised publicly available data, the sources for which were cited in the study, and as such ethical approval was not required

## Informed Consent Statement

The study used only anonymised publicly available data, and no human or animal subjects were recruited for the study.

## Data Availability Statement

The data used in the study are publicly available and are cited in the manuscript.

## Acknowledgments

The authors extend their thanks to the surveillance team at VetStat and The Danish Integrated Antimicrobial Resistance Monitoring and Research Programme (DANMAP) for curating the data used in the study and for making them publicly available.

## Appendix

**Appendix 1.** classification of antibiotics in this study.

| Listed in human resistance dataset | Unified class | Listed in animal use dataset | Unified class |
| --- | --- | --- | --- |
| Amikacin | Aminoglycosides | Aminoglycosides | Aminoglycosides |
| Amoxicillin/Clavulanic acid | Penicillins | Amphenicols | Other |
| Ampicillin | Penicillins | Cephalosporins | Cephalosporins |
| Apramycin | Aminoglycosides | Fluoroquinolones | Fluoroquinolones |
| Azithromycin | Macrolides | Lincosamides | Lincosamides |
| Cefotaxime | Cephalosporins | Macrolides | Macrolides |
| Ceftazidime | Cephalosporins | Other | Other |
| Ceftiofur | Cephalosporins | Penicillins (ext.) | Penicillins |
| Chloramphenicol | Other | Penicillins (sim.) | Penicillins |
| Ciprofloxacin | Fluoroquinolones | Quinolones | Quinolones |
| Colistin | Polymyxins | Sulfonamides/Trimethoprim | Sulfonamides |
| Ertapenem | Carbapenems | Tetracyclines | Tetracyclines |
| Erythromycin | Macrolides | Tiamulines | Other |
| Florfenicol | Amphenicols |  |  |
| Gentamicin | Aminoglycosides |  |  |
| Meropenem | Carbapenems |  |  |
| Nalidixic acid | Quinolones |  |  |
| Neomycin | Aminoglycosides |  |  |
| Spectinomycin | Aminoglycosides |  |  |
| Streptomycin | Aminoglycosides |  |  |
| Sulfonamide | Sulfonamide |  |  |
| Tetracycline | Tetracycline |  |  |
| Tigecycline | Others |  |  |
| Trimethoprim | Others |  |  |

**Appendix 2.**
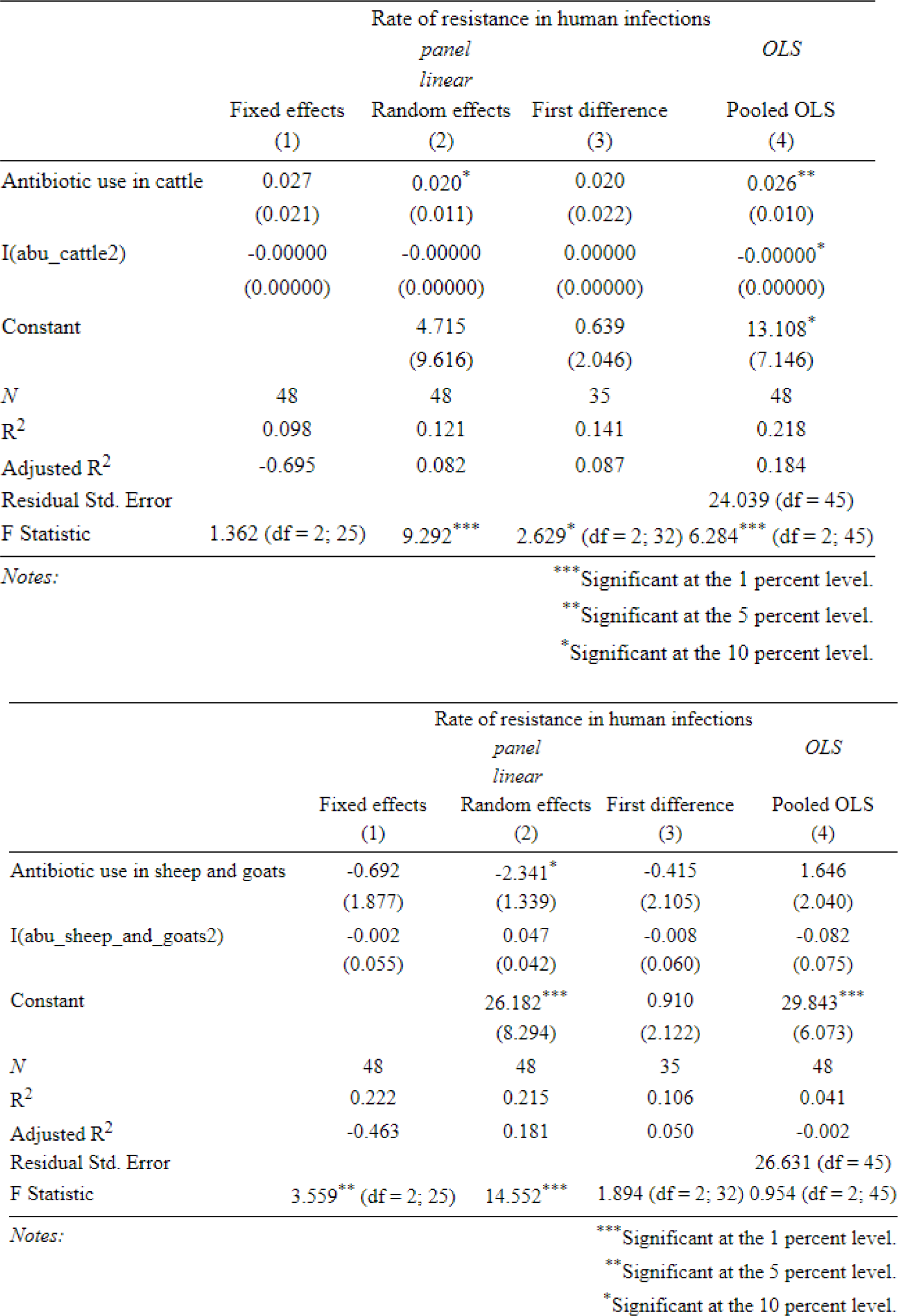
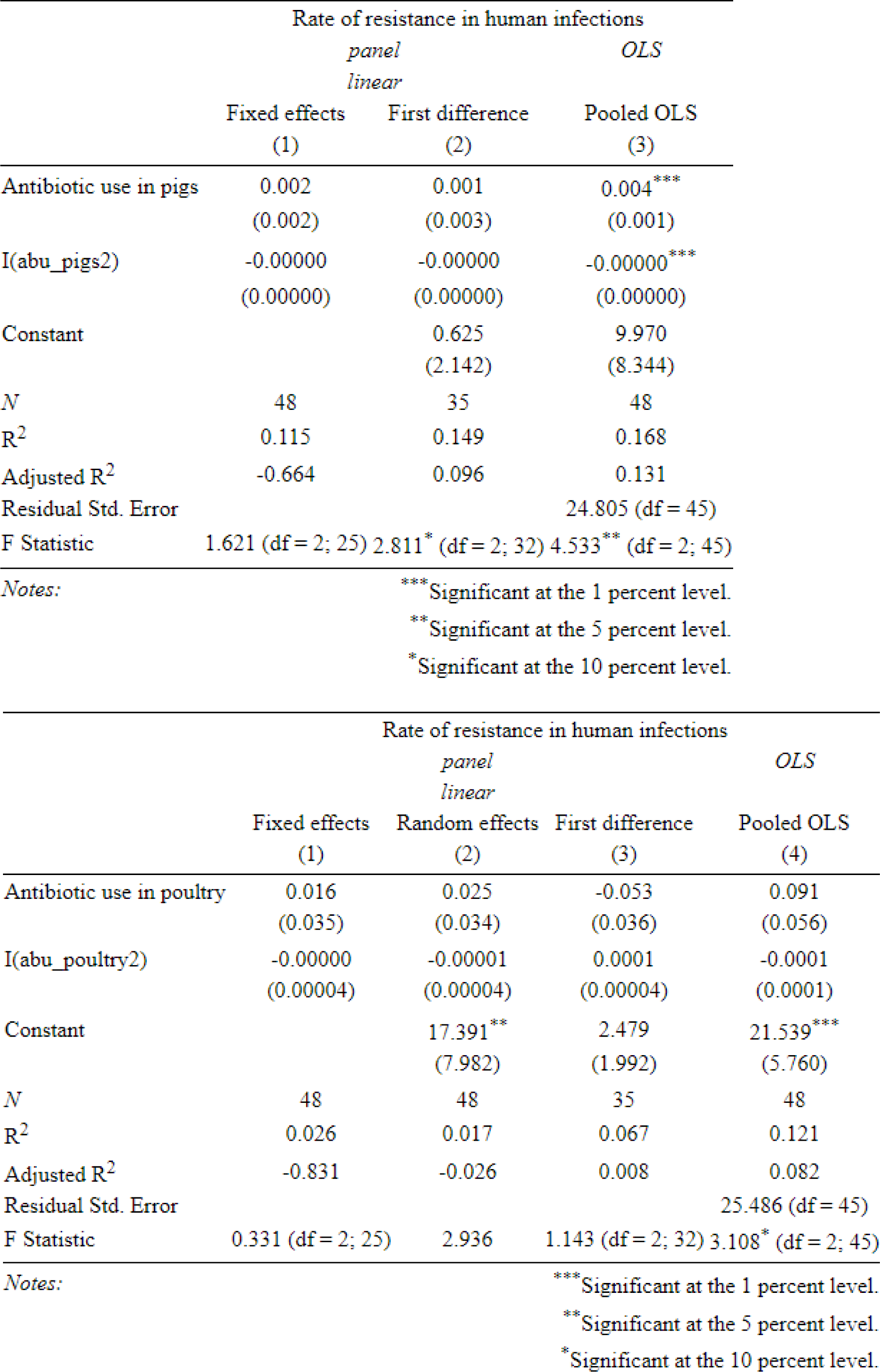
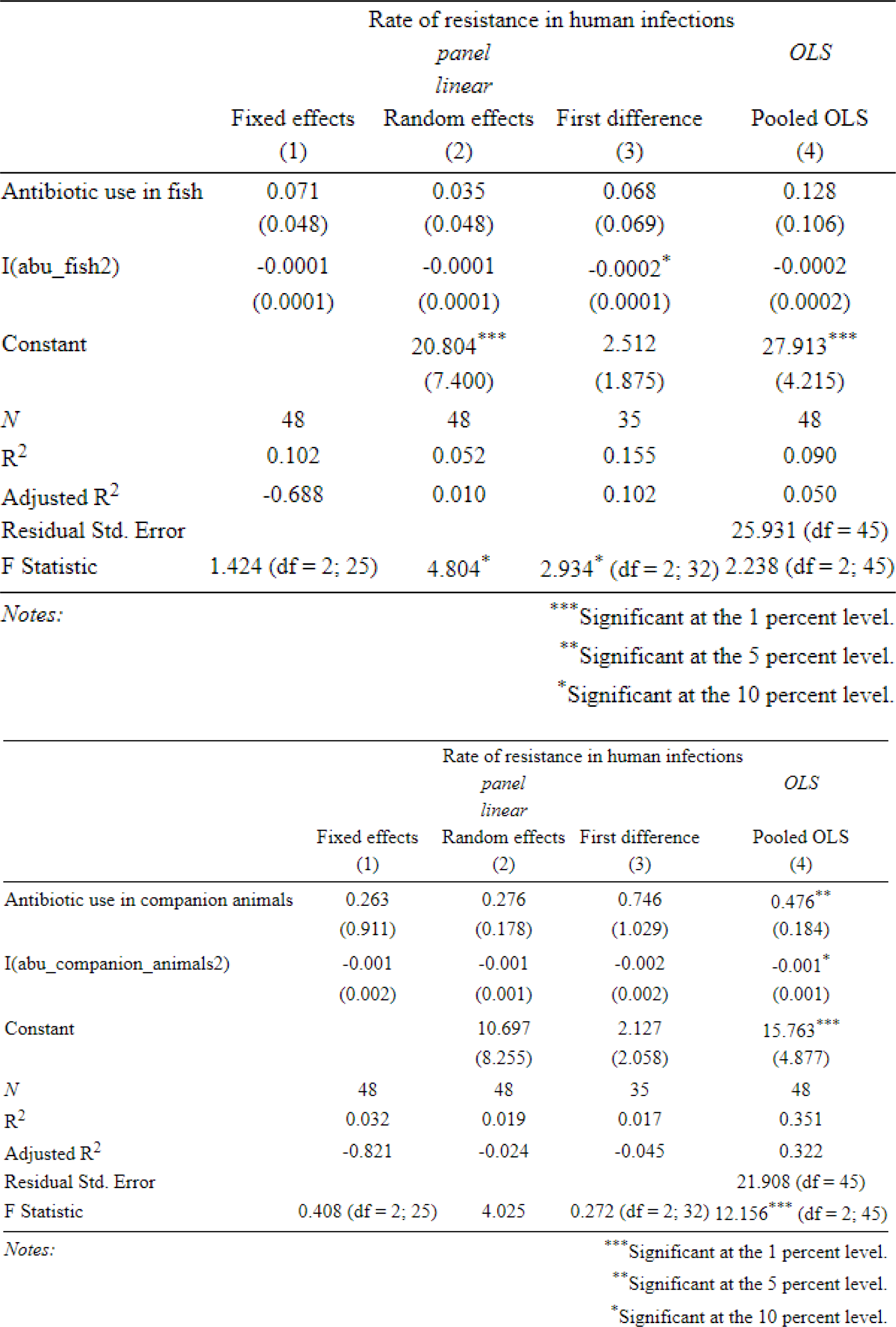
quadratic specifications.

**Appendix 3.** Specifications with additional covariates.

|  | Rate of resistance in human infections |  |  |  |
| --- | --- | --- | --- | --- |
|  | *panel* |  |  | *OLS* |
|  | *linear* |  |  |  |
|  | Fixed effects<br>(1) | Random effects<br>(2) | First difference<br>(3) | Pooled OLS<br>(4) |
| Antibiotic use in cattle | -0.001<br>(0.029) | 0.007***<br>(0.002) | 0.002<br>(0.002) | 0.007**<br>(0.002) |
| Cattle population |  |  |  | 0.00002<br>(0.0002) |
| GDP per capita, PPP |  | 0.002***<br>(0.001) |  | 0.002**<br>(0.001) |
| Constant |  | -67.061**<br>(34.165) | 3.577<br>(5.595) | -106.690<br>(296.557) |
| *N* | 48 | 48 | 39 | 48 |
| *R*<sup>2</sup> | 0.00005 | 0.270 | 0.022 | 0.271 |
| Adjusted *R*<sup>2</sup> | -0.424 | 0.238 | -0.005 | 0.221 |
| Residual Std. Error |  |  |  | 23.484 (df = 44) |
| F Statistic | 0.001 (df = 1; 33) | 16.667*** | 0.814 (df = 1; 37) | 5.441*** (df = 3; 44) |
| *Notes:* |  |  |  | *** Significant at the 1 percent level.<br>** Significant at the 5 percent level.<br>* Significant at the 10 percent level. |
|  | Rate of resistance in human infections |  |  |  |
|  | *panel* |  |  | *OLS* |
|  | *linear* |  |  |  |
|  | Fixed effects<br>(1) | Random effects<br>(2) | First difference<br>(3) | Pooled OLS<br>(4) |
| Antibiotic use in sheep and goats | -0.382<br>(0.828) | -0.026<br>(0.611) | -0.856<br>(1.254) | -0.026<br>(0.611) |
| Sheep population |  | -0.00005<br>(0.001) |  | -0.00005<br>(0.001) |
| GDP per capita, PPP |  | 0.002**<br>(0.001) |  | 0.002**<br>(0.001) |
| Constant |  | -62.394<br>(122.152) | 5.319<br>(5.943) | -62.394<br>(122.152) |
| *N* | 48 | 48 | 39 | 48 |
| *R*<sup>2</sup> | 0.006 | 0.149 | 0.012 | 0.149 |
| Adjusted *R*<sup>2</sup> | -0.415 | 0.091 | -0.014 | 0.091 |
| Residual Std. Error |  |  |  | 25.368 (df = 44) |
| F Statistic | 0.212 (df = 1; 33) | 7.693* | 0.465 (df = 1; 37) | 2.564* (df = 3; 44) |
| *Notes:* |  |  |  | *** Significant at the 1 percent level.<br>** Significant at the 5 percent level.<br>* Significant at the 10 percent level. |
|  | *panel* |  |  | *OLS* |
|  | *linear* |  |  |  |
|  | Fixed effects<br>(1) | Random effects<br>(2) | First difference<br>(3) | Pooled OLS<br>(4) |
| Antibiotic use in pigs | -0.0001<br>(0.001) | 0.001<br>(0.0003) | -0.0001<br>(0.001) | 0.001<br>(0.0003) |
| Pig population |  |  |  | -0.00001<br>(0.00001) |
| GDP per capita, PPP |  | 0.002***<br>(0.001) |  | 0.002**<br>(0.001) |
| Constant |  | -84.444**<br>(36.916) | 4.447<br>(6.009) | 66.663<br>(157.153) |
| *N* | 48 | 48 | 39 | 48 |
| *R*<sup>2</sup> | 0.0003 | 0.193 | 0.001 | 0.211 |
| Adjusted *R*<sup>2</sup> | -0.424 | 0.157 | -0.026 | 0.157 |
| Residual Std. Error |  |  |  | 24.428 (df = 44) |
| F Statistic | 0.009 (df = 1; 33) | 10.776*** | 0.052 (df = 1; 37) | 3.916** (df = 3; 44) |
| *Notes:* |  |  | *** Significant at the 1 percent level. |  |
|  |  |  | ** Significant at the 5 percent level. |  |
|  |  |  | * Significant at the 10 percent level. |  |
|  | *panel* |  |  | *OLS* |
|  | *linear* |  |  |  |
|  | Fixed effects<br>(1) | Random effects<br>(2) | First difference<br>(3) | Pooled OLS<br>(4) |
| Antibiotic use in poultry | -0.016<br>(0.039) | 0.034**<br>(0.016) | -0.019<br>(0.026) | 0.017<br>(0.017) |
| Chicken population |  | -0.00000<br>(0.00000) |  | -0.00000<br>(0.00000) |
| GDP per capita, PPP |  |  |  | 0.002**<br>(0.001) |
| Constant |  | 29.105***<br>(10.290) | 5.398<br>(5.925) | -62.130<br>(38.891) |
| *N* | 48 | 48 | 39 | 48 |
| *R*<sup>2</sup> | 0.005 | 0.117 | 0.014 | 0.210 |
| Adjusted *R*<sup>2</sup> | -0.417 | 0.078 | -0.012 | 0.156 |
| Residual Std. Error |  |  |  | 24.439 (df = 44) |
| F Statistic | 0.161 (df = 1; 33) | 4.553 | 0.543 (df = 1; 37) | 3.900** (df = 3; 44) |
| *Notes:* |  |  | *** Significant at the 1 percent level. |  |
|  |  |  | ** Significant at the 5 percent level. |  |
|  |  |  | * Significant at the 10 percent level. |  |
|  | *panel* |  |  | *OLS* |
|  | *linear* |  |  |  |
|  | Fixed effects<br>(1) | Random effects<br>(2) | First difference<br>(3) | Pooled OLS<br>(4) |
| Antibiotic use in fish | -187.655<br>(597.449) | 0.064***<br>(0.023) | 0.624<br>(0.942) | 1.104<br>(0.963) |
| Fisheries production |  |  |  | 0.003<br>(0.004) |
| GDP per capita, PPP |  | 0.003***<br>(0.001) |  |  |
| Constant |  | -92.557***<br>(34.911) | 9.924<br>(15.235) | -64.850<br>(138.440) |
| *N* | 9 | 48 | 7 | 9 |
| *R*<sup>2</sup> | 0.032 | 0.275 | 0.081 | 0.214 |
| Adjusted *R*<sup>2</sup> | -1.582 | 0.242 | -0.103 | -0.049 |
| Residual Std. Error |  |  |  | 26.789 (df = 6) |
| F Statistic | 0.099 (df = 1; 3) | 17.033*** | 0.438 (df = 1; 5) | 0.814 (df = 2; 6) |

Notes:
\*\*\* Significant at the 1 percent level.
\*\* Significant at the 5 percent level.
\* Significant at the 10 percent level.

|  | Rate of resistance in human infections |  |  |  |
| --- | --- | --- | --- | --- |
|  | *panel* |  |  | *OLS* |
|  | *linear* |  |  |  |
|  | Fixed effects<br>(1) | Random effects<br>(2) | First difference<br>(3) | Pooled OLS<br>(4) |
| Antibiotic use in companion animals | 0.225<br>(0.304) | 0.156***<br>(0.039) | 0.165***<br>(0.042) | 0.156***<br>(0.039) |
| Pet ownership |  | -214.733<br>(257.027) |  | -214.733<br>(257.027) |
| GDP per capita, PPP |  | 0.004<br>(0.003) |  | 0.004<br>(0.003) |
| Constant |  | -72.676*<br>(38.268) | 7.437<br>(5.690) | -72.676*<br>(38.268) |
| *N* | 31 | 31 | 25 | 31 |
| *R*<sup>2</sup> | 0.028 | 0.497 | 0.403 | 0.497 |
| Adjusted *R*<sup>2</sup> | -0.535 | 0.441 | 0.377 | 0.441 |
| Residual Std. Error |  |  |  | 21.447 (df = 27) |
| F Statistic | 0.545 (df = 1; 19) | 26.662*** | 15.513*** (df = 1; 23) | 8.887*** (df = 3; 27) |

Notes:
\*\*\* Significant at the 1 percent level.
\*\* Significant at the 5 percent level.
\* Significant at the 10 percent level.

**Appendix 4.**
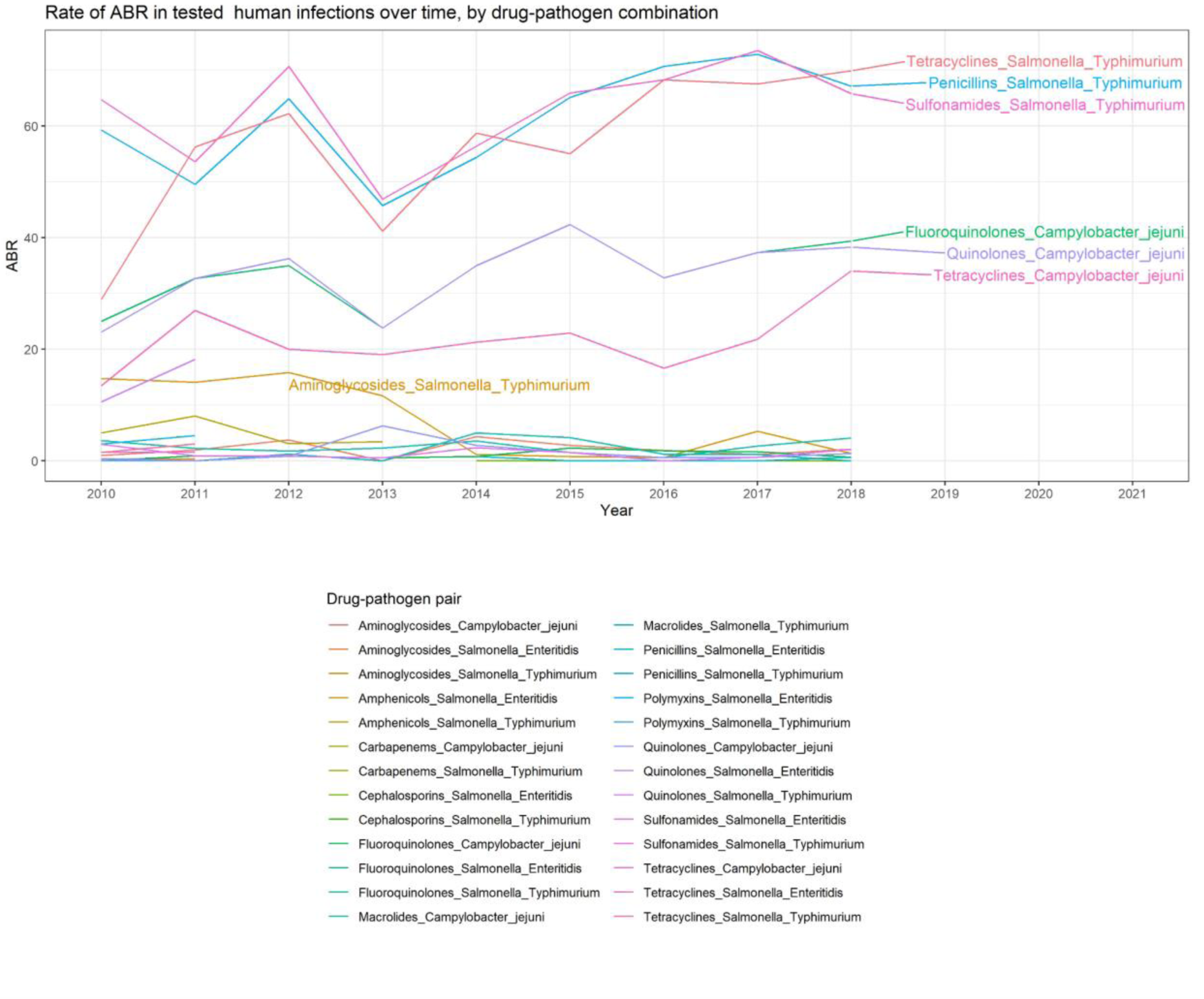
Figure 3 with full legend.

## Footnotes

1 One Health refers to the interplay between human, animal and environmental health

